# An Improved Pipeline for Constructing UK Biobank Brain Imaging Confounds

**DOI:** 10.1101/2025.11.21.25340740

**Authors:** Lav Radosavljević, Thomas Maullin-Sapey, Fidel Alfaro-Almagro, Paul McCarthy, Thomas E. Nichols, Stephen M. Smith

**Affiliations:** Nuffield Department of Population Health, University of Oxford; School of Mathematics, University of Bristol; Nuffield Department of Clinical Neurosciences, University of Oxford

## Abstract

UK Biobank (UKB) brain imaging data is a one-of-a-kind resource for studying the links between the brain and demographic-, lifestyle- and genetic data. When establishing such links, it is crucial to account for confounding effects caused by the acquisition of fMRI images, as well as demographic confounding factors. UKB brain imaging confounds are constructed through variable selection by the proportion of variance explained in the Imaging Derived Phenotypes (IDPs), from tens of thousands of possible confounds. The current implementation of this pipeline is very computationally intensive and has a large memory footprint, largely due to the varying patterns of missing data in IDPs. This makes it impractical for many users of UK Biobank brain imaging data. We propose a fast and memory efficient multivariate pipeline for constructing imaging confounds using mean imputation combined with a bias-corrected estimator of *R*^2^, the proportion of confound variance explained in an IDP. Building on this, we also improve the pipeline in order to better select confounds that explain unique variance in IDPs, and non-imaging variables of interest, so called nIDPs. The new implementation leads to a more compact set of confounds that explains roughly the same amount of variance, and runs in around 1 hour on a single CPU.

## 1 Introduction

The UK Biobank (UKB) brain imaging analysis pipeline (Alfaro-Almagro et al., 2018) produces around 6,000 Imaging Derived Phenotypes (IDPs), and also over 10, 000 Quality Control (QC) and diagnostic variables that may be valuable as confounds when estimating associations between IDPs and other outcome variables (nIDPs, non-imaging-derived phenotypes), such as lifestyle variables, health outcomes and genetics. In previous work (Alfaro-Almagro et al., 2021) we systematically identified around 1,000 confounds that explained non-trivial variance in IDP on nIDP associations. Our previous approach to identifying UKB imaging confounds selects confounds, one-by-one, that explain a high proportion of variance across all IDPs and/or a very high proportion of variance in individual IDPs. As this is a serial and not joint process, it has the disadvantage of selecting several highly correlated confounds, leading to redundancy in the confound set. To avoid redundancy in the selected confounds we propose using a forward stepwise selection procedure, at each step selecting the next confound variable that increases *R*^2^ (proportion of variance explained) the most over IDPs. However, with varying missingness (different IDPs have different patterns of missing or outlier values), stepwise regression over thousands of IDPs is computationally burdensome, due to the need to fit a unique linear model for each IDP. We solve this via mean imputation of the IDPs and post-imputation correction of all *R*^2^-estimates. We show that this does not degrade the choice of confounds, and that we can estimate a largely equivalent but much more compact set of brain imaging confounds.

## 2 Background

Handling confounding effects is a ubiquitous problem in epidemiological and biostatistical research (Basto et al., 2023, Brenner and Blettner, 1997, Greenland and Morgenstern, 2001, Morabia, 2011, Robins and Morgenstern, 1987). In classical epidemiological modelling, sex, age, socio-economic status and smoking status are typical examples of confounding variables that modify the relationship between exposure and outcome (Hajat et al., 2021, McNamee, 2003, Papageorgiou, 2019). In the case of imaging variables, the situation is much more complicated, as the imaging process itself causes nuisance effects that are associated with brain IDPs and nIDPs. UKB brain imaging confounds are constructed by initially considering a relatively small set of imaging confounds such as sex, age, site, head-motion, scanner-protocol, table position, etc., and treating their effect on IDPs as linear and additive. To fully capture the complicated relationships between confounds and IDPs, non-linear transformations of these confounds along with interaction terms and time trends are also considered. However, since the number of possible confounds is in the tens of thousands, a subset of the most relevant potential confounds is selected. In particular, a subset of nonlinear confounds are first selected based on the variance explained in the IDPs (after first regressing out linear confounds); then a subset of variable interactions are selected in the same way. Finally, a set of confounds is constructed to capture trends caused by scanner drift over time or distribution shifts in the population, as well as Time of Day (ToD) trends, using the following procedure: All confounds constructed so far are regressed out of the IDPs, and the residuals are ordered by ToD and Date, separately, are smoothed, and the Principal Components (PCs) of these two smoothed matrices are selected such that they explain 90% of the variance of the smoothed data. These PCs form the basis of the time trends confounds in the IDPs. Currently, the whole pipeline yields 966 brain imaging confounds.

Previously, because the evaluation of potential confounds was computationally expensive, each potential confound was considered for inclusion in parallel. As this can lead to mass redundancy, it would be preferable to use stepwise forward selection, building up a set of the most important confounds, and only adding additional potential confounds to this set if they explain IDP variance that is unique compared with the existing working confound set. The main obstacle to forward selection in this pipeline is heterogeneous, i.e., variable-specific missingness in the IDPs. Specifically, if we insist on using Complete Cases (CC), each IDP has a unique design matrix associated with it, and therefore we require a unique linear model for each IDP. Since there are around 6,000 IDPs, it is very computationally burdensome to perform forward selection over all of them simultaneously. A classical solution to this problem is imputation of missing data, but this biases the selection process, because the effect of imputation on *R*^2^, i.e., the variance explained, varies between IDPs depending on their Signal to Noise Ratio (SNR) and their rate of missingness. For example, if we use an imputation method that shrinks *R*^2^ the higher the rate of missingness, the IDPs with low missingness consequently disproportionally influence the selection process, thus biasing all downstream analysis using confounds.

## 3 Methods

For a scalar random variable *y* following a linear model of the form

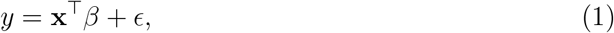

where **x** is a multivariate random vector, *β* a fixed set of coefficients and *ϵ* a mean zero random variable independent of **x**, *R*^2^ is defined as

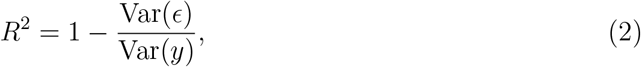

i.e., the proportion of variance in *y* that can be explained by **x** (Fisher, 1928, Olkin and Pratt, 1958). For a linear model fitted on data from independent draws of **x** and *y*, the standard point estimate of *R*^2^ is

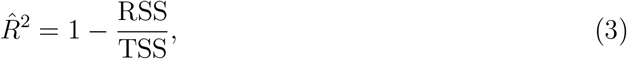

where RSS is the residual sum of squares and TSS is the total sum of squares. This is a consistent estimator of *R*^2^ (Koerts and Abrahamse, 1969). Using the change in *R*^2^ as the criterion for variable selection is a common modelling choice in forward selection pipelines (Blanchet et al., 2008, Ghani and Ahmad, 2010, James et al., 2023, Lindsey and Sheather, 2010). However, we cannot use the standard estimator of *R*^2^ for this purpose if the missing data of the dependent variable *y* has been imputed, as this estimator is no longer consistent.

### 3.1 Correcting *R*^2^ after mean imputation

To solve the problem of *R*^2^ bias under imputation, we propose a simple correction of the *R*^2^ estimate following mean imputation. If 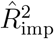 is the standard *R*^2^-estimate calculated on mean imputed data, our proposed corrected estimate is

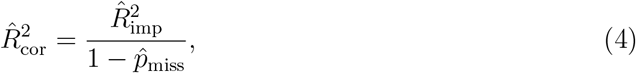

where 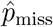 is the rate of missingness in our dependent variable, i.e., our IDP. What this means intuitively is that if the rate of missingness in the dependent variable is *p*_miss_, mean imputation leads to a change in *R*^2^ by a factor of 1 − *p*_miss_. In the Supplementary Material, we prove that Eqn. (4) is a consistent estimator of the true *R*^2^ under Missing Completely at Random (MCAR) (see Section S1.1).

### 3.2 Proposed Forward Selection Method

Our forward selection method assumes the following linear model for each IDP **y**:

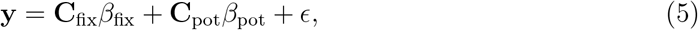

where **C**_fix_ is the matrix of fixed confounds (e.g. age, sex, or previously selected confounds), and **C**_pot_ is the matrix of potential confounds from which we want to select. We assume that many of the potential confounds are redundant, that missing data in the IDPs occurs under MCAR, and that there is no missing data in the confounds. Figure 1 illustrates this selection problem. Using our corrected estimate, we select a maximum of two variables to add to our confound set in each iteration: the variable that has the maximal change in corrected 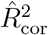 for a single variable, and the one that has the maximal average change in 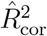 over all variables. We set two pre-determined thresholds *τ*_max_ and *τ*_mean_, determining when to not include variables based on the two criteria. The algorithm stops when the maximal change in 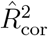 is lower than *τ*_max_ and the maximal average change is lower than *τ*_mean_. The default values we use for these thresholds are *τ*_max_ = 0.0025 and *τ*_mean_ = 0.0005. If the two selected variables are too highly correlated with each other (having an absolute Pearson correlation exceeding 0.95), we only include the variable selected by the mean criterion. An essential feature of using mean imputation is that our algorithm can be implemented in a vectorised fashion, such that each iteration only consists of matrix manipulations; pseudocode can be found in the Supplementary Material (Section S1.2), and python code is openly available at https://git.fmrib.ox.ac.uk/fsl/fastconfounds/.

**Figure 1.**
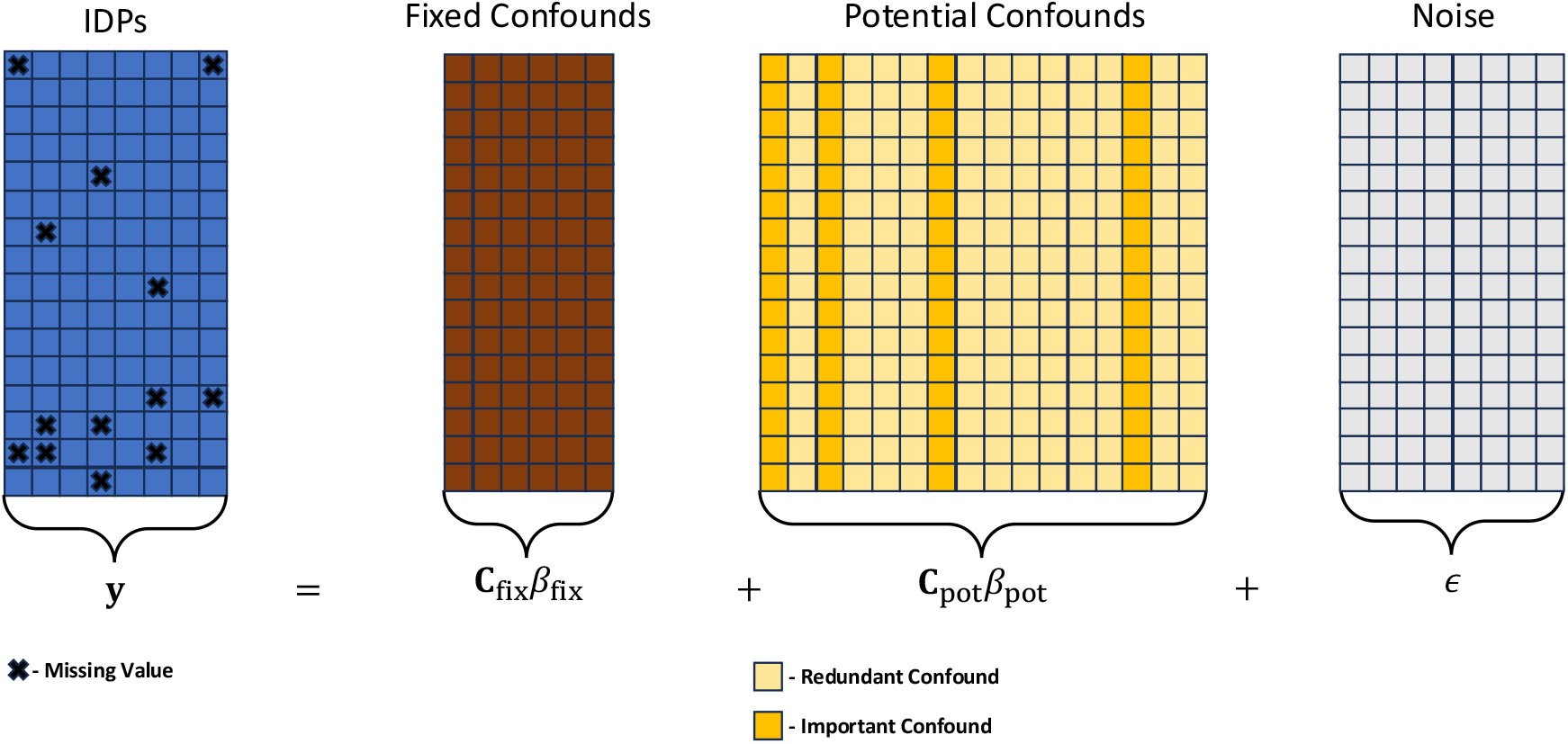
Illustration of the regression equation that is the focus of this work. We assume that only a very small subset of potential confounds are relevant to the IDPs and that there is heterogeneous missingness in the IDPs.

### 3.3 Double Selection

In our previous pipeline for constructing imaging confounds, we only select confounds in the context of their relationship to IDPs. However, since confounds can relate to both IDPs and nIDPs, it makes sense to also consider nIDPs when constructing imaging confounds. So-called *double selection*, i.e., selecting all variables that are predictive of either outcome or exposure, has been shown to have desirable properties in terms of false positive control and power when controlling for confounding variables in high-dimensional settings (Belloni et al., 2014). We therefore include 979 nIDPs in the selection process. The nIDPs were chosen by excluding all nIDPs with rates of missingness higher than 20% and all binary nIDPs where the most common value represents over 90% of cases. We also consider a version of the pipeline with *single selection*, i.e., where only IDPs are used for selection, and compare it to the version using double selection.

### 3.4 Proposed Pipeline

Using the corrected *R*^2^ estimate, we construct a forward selection pipeline for constructing confounds that closely follows the previous pipeline (Alfaro-Almagro et al., 2021), while applying forward selection instead of univariate selection. The pipeline follows this procedure:

1. We construct “linear confounds”, which are the raw continuous confounds and categorical confounds encoded as dummy variables.
2. We construct “non-linear confounds”, three sets of transformation of all continuous linear confounds. There is a squaring of each variable (after centering); a quantile-transform to a standard normal distribution (Peterson and Cavanaugh, 2019); and a squaring of the normal-transformed variables.
3. We construct a baseline set of confounds consisting of age, sex and site, and select a subset of the remaining linear and non-linear variables using forward selection.
4. We construct “crossed terms”, considering all possible variable interactions for the selected variables (i.e., second-order multiplicative interactions between all the above confounds) and again perform forward selection to select a subset.
5. To construct time trends, we mean-impute the IDPs and regress out the selected confounds.
  a. For each site, we perform Gaussian kernel smoothing for all the IDPs’ residuals over time of day. We perform PCA on the smoothed residuals and include enough PCs to explain 99% of the variance.
  b. Using all extracted PCs, we again perform forward selection to obtain a subset of PCs.
  c. We regress the newly selected variables out of the IDPs and perform smoothing over date. We extract PCs again and perform forward selection.

Flowcharts illustrating the old and new pipelines are shown in Figures 2 and 3. Confounds are constructed using the full ≈ 63, 000 subjects in the UK Biobank brain imaging cohort. Almost all IDPs are used in the pipeline, but 50 ASL IDPs are currently excluded for having too high rates of missingness, with some of them having over 90% missing data. For the new pipeline, double-selection is performed by adding the nIDPs to the IDP data, with the only exception being the construction of time trends, as nIDPs and IDPs do not have the same acquisition times.

**Figure 2.**
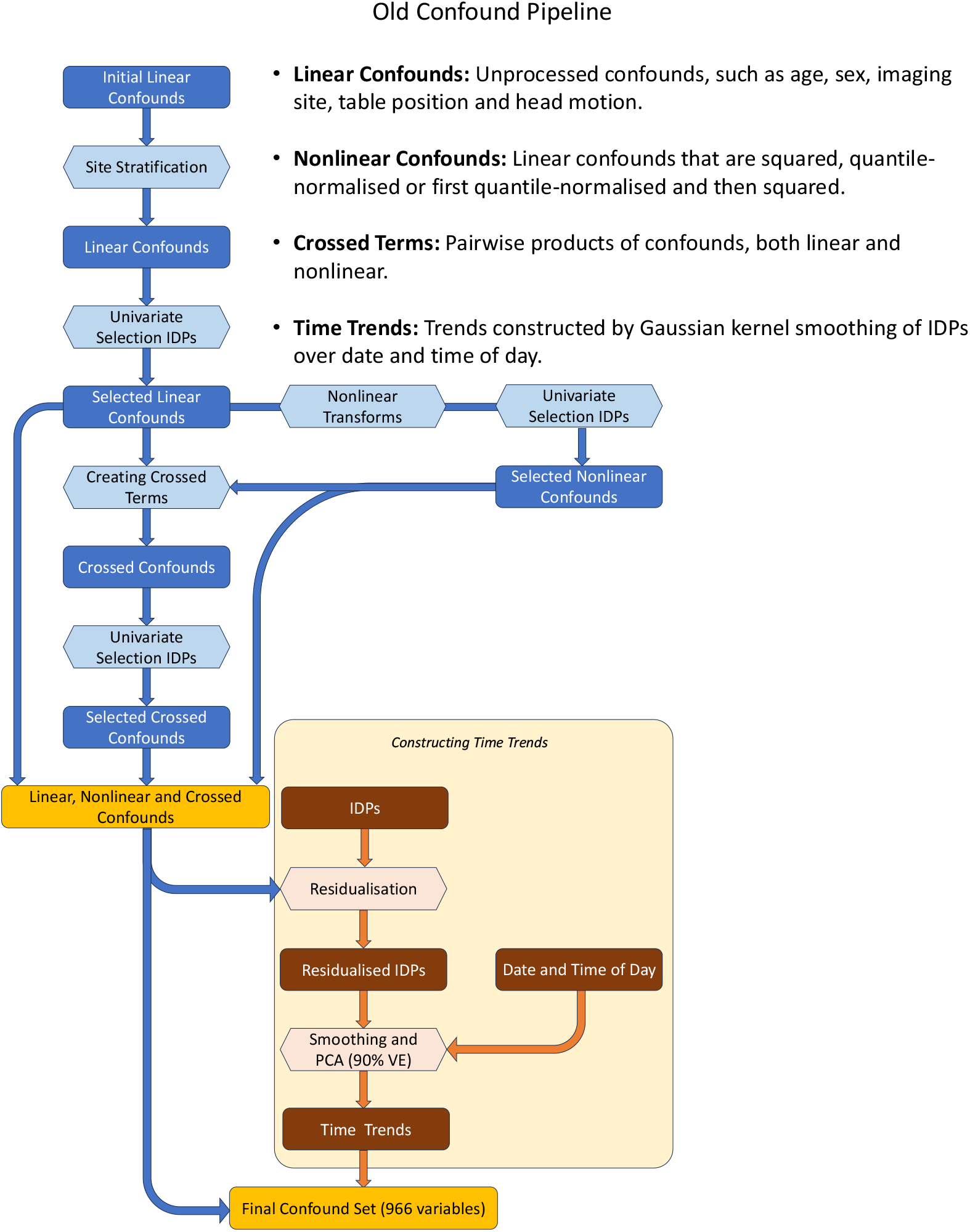
Flow chart illustrating the old confound pipeline. Unlike the new pipeline, the old pipeline uses univariate selection to include confounds, instead of multivariate forward selection. Another difference is in constructing time trends; the old pipeline includes all PCs up to 90% variance explained, regardless of their ability to explain IDPs, whereas the new pipeline performs forward selection to choose PCs. Only IDPs are used for selection, whereas the new pipeline uses nIDPs as well

**Figure 3.**
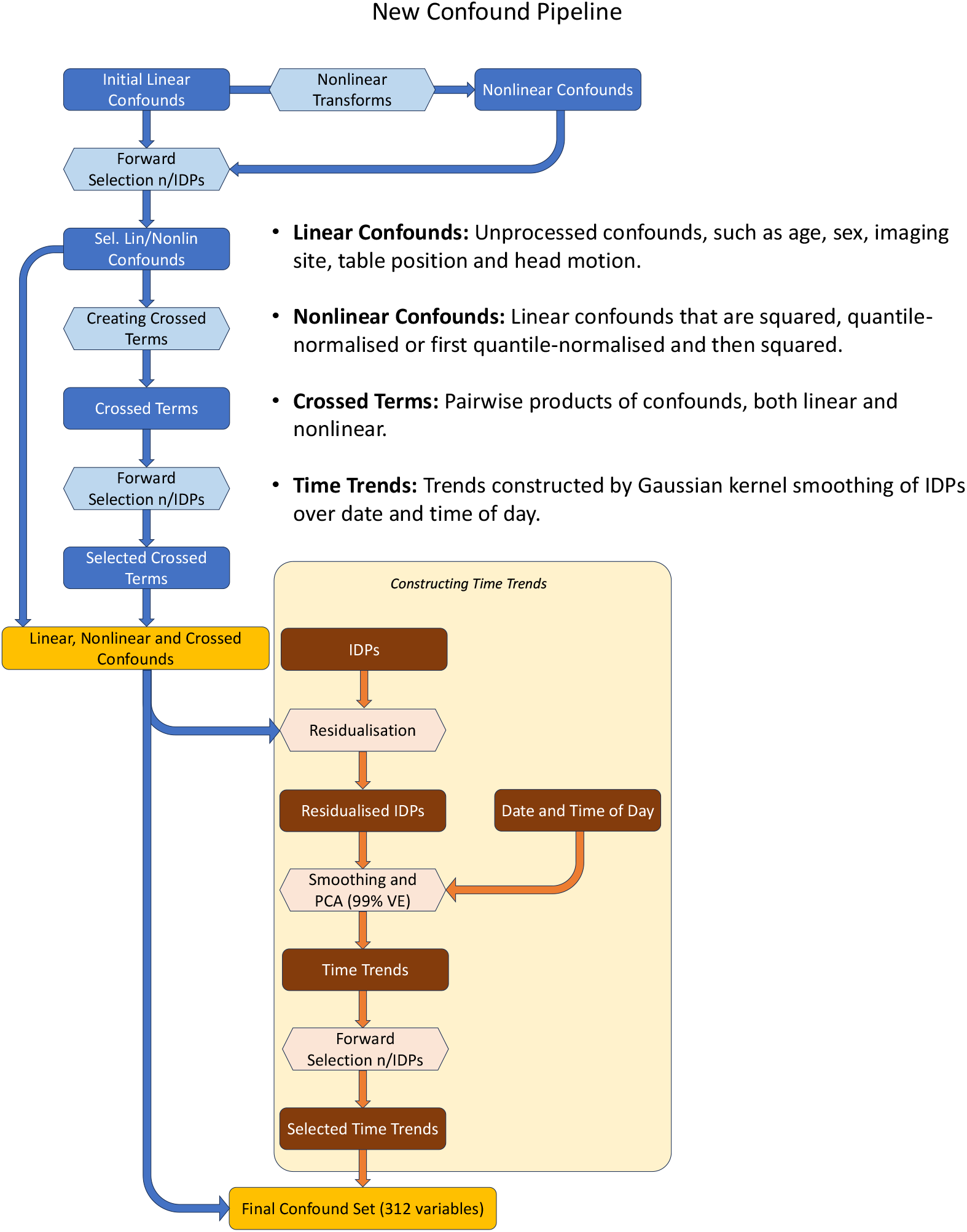
Flow chart illustrating the new confound pipeline. Unlike the old pipeline, the new pipeline uses multivariate forward selection to include confounds, instead of univariate selection. Another difference is in constructing time trends; the old pipeline includes all PCs up to 90% variance explained, regardless of their ability to explain IDPs, whereas the new pipeline performs forward selection to choose PCs. The new pipeline uses nIDPs in addition to IDPs for selection.

### 3.5 Evaluating the New Pipeline

We evaluate the new pipeline relative to the old pipeline in a number of ways. First we evaluate the corrected estimator 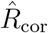 by comparing it to 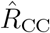, the standard estimator of *R*^2^ using CC (i.e., the gold standard under MCAR using complete cases data only). We make this comparison over all IDPs and our subset of nIDPs as part of double-selection (see Section 3.3). To assess whether the two confound sets explain the same amount of variance in each variable, we compare the variance explained by the old and new confound sets in a Bland-Altman plot. Additionally, we compare the actual variance explained, not just the amount. We assess this by measuring the increase in variance explained by concatenating the two confound sets; if the same confound variance is explained, there will be no increase. We also examine the correlation of each IDPs/nIDPs after deconfounding using old vs. new confounds; a high correlation indicates that deconfounding using old and new confounds has a roughly equivalent effect.

Lastly, we compare the p-values for univariate associations between IDPs and nIDPs, when using old vs. new confounds for deconfounding. Here we also construct a Bland-Altman plot showing the agreement in − log p-values when using old and new confounds for deconfounding.

When comparing the variance explained in the IDPs/nIDPs between old and new confound sets, we use adjusted *R*^2^ (Ezekiel, 1930) instead of the regular estimate, to account for the fact that there are different numbers of confounds in the two sets:

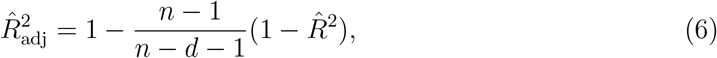

where *n* is the number of subjects, *d* the number of confounds and 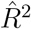 the standard 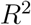 estimator.

## 4 Results

Our new pipeline yields 312 confounds, compared to 966 confounds for the old pipeline. The lower number of confounds is a result of forward selection in the new pipeline, which avoids the joint selection of highly correlated variables caused by greedy univariate selection in the old pipeline. Table 1 shows the number of confounds in each confound group for the old and new pipelines. The most stark difference is in the time trend confounds, where the new pipeline only constructs two confounds compared to 186 for the old pipeline. The reason for this is that in the old pipeline, after smoothing the residualised IDPs all PCs up to 90% total variance explained are included, whereas in the new pipeline, the same forward selection procedure is applied to the extracted PCs and only PCs that pass the thresholds for variance explained are included.

**Table 1.** Number of confounds in each confound group, for old and new confounds. We see that the new pipeline constructs significantly fewer confounds. The difference is particularly stark for the time trend confounds. This is because the new pipeline further reduces them by applying forward selection to the PCs that are extracted from the smoothed IDP residuals.

|  | Old Pipeline | New Pipeline |
| --- | --- | --- |
| **Linear** | 378 | 64 |
| **Non-Linear** | 178 | 52 |
| **Crossed Terms** | 224 | 194 |
| **Time Trends** | 186 | 2 |
| **Total** | **966** | **312** |

Figure 4 shows the relative difference between 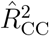 and 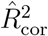 plotted against the rate of missingness, for IDPs and nIDPs separately. The percentage of relative difference here means:

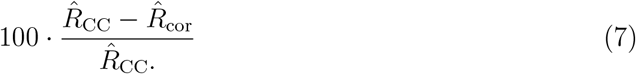

**Figure 4.**
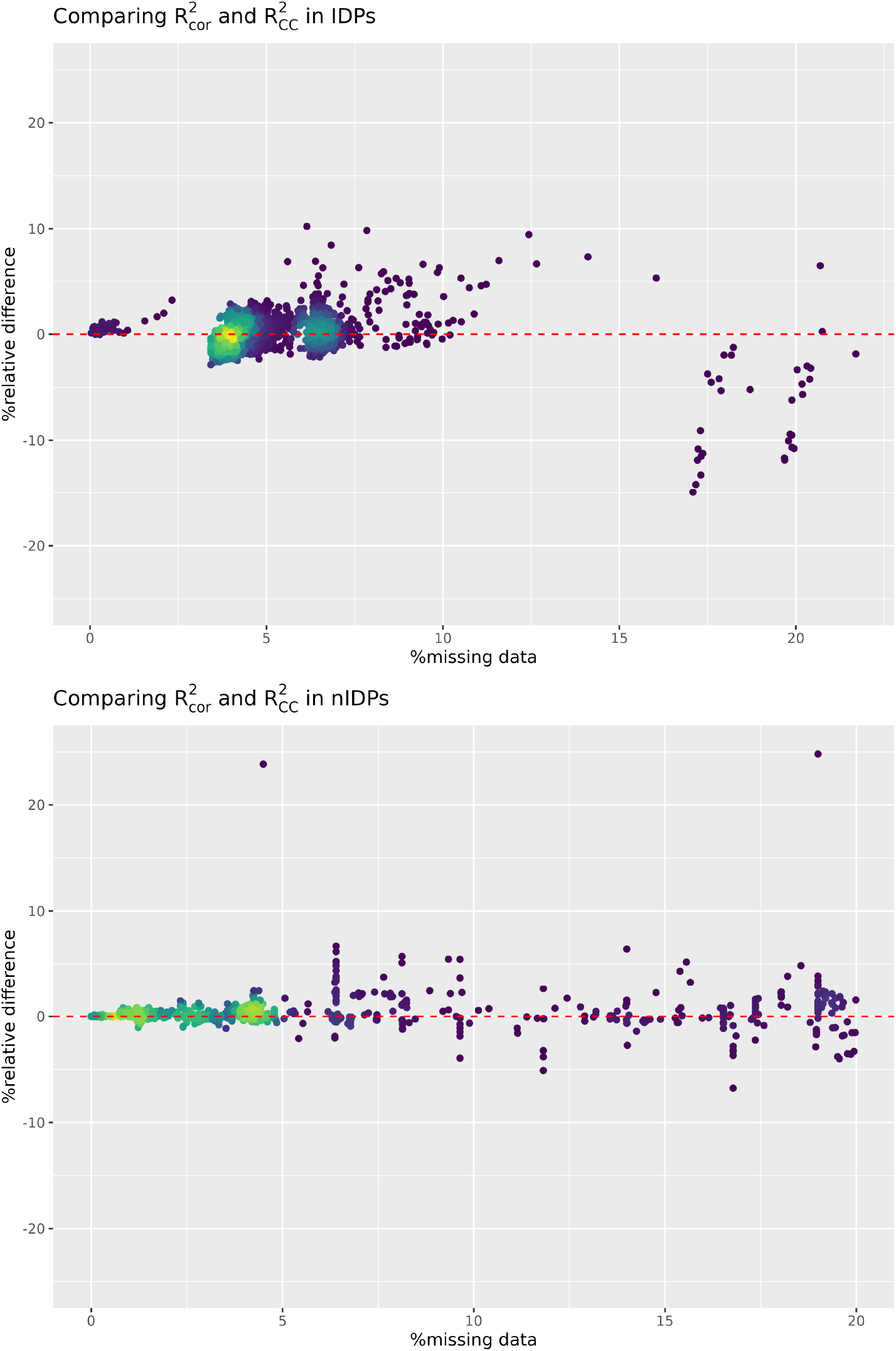
Scatter plots of missingness rate against the percentage difference between 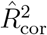 and 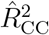. For the vast majority of IDPs and nIDPs, the two estimates are within 5% relative difference of each other (99.2% of IDPs and 98.6% of nIDPs), which is sufficiently close for our purposes.

Each point in the plot represents a single IDP/nIDP and compares the estimated proportion of variance explained in the variable by the old confound set, as calculated using 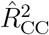 and 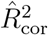, plotted as percent relative difference. There is strong agreement between the two estimators at different levels of missingness, with the vast majority of estimates being within 5% relative difference of each other (99.2% for IDPs and 98.6% for nIDPs). No estimates differ more than 25% from each other.

Figure 5 shows the average proportion of variance explained in the IDPs and nIDPs by the old and new confound sets, as well as the proportion explained by the union of both confound sets. The two confound sets explain similar proportions of variance on average, but the variance explained is distributed differently between confound groups. In particular, the non-linear confounds explain a larger portion of variance in the new confound sets. This is expected, as the non-linear and linear confounds are selected simultaneously in the new confound pipeline, while the linear are selected before the non-linear in the old pipeline, thereby leading to a larger proportion of variance initially accounted for by linear confounds. Additionally, we see that the crossed terms explain relatively more variance in the new confound set, and the time trends explain relatively less variance for the new confound set. Importantly, there is no notable increase in variance explained when combining new and old confounds, meaning that almost all important mutual variance between confounds and IDPs/nIDPs is captured by the new confound set.

**Figure 5.**
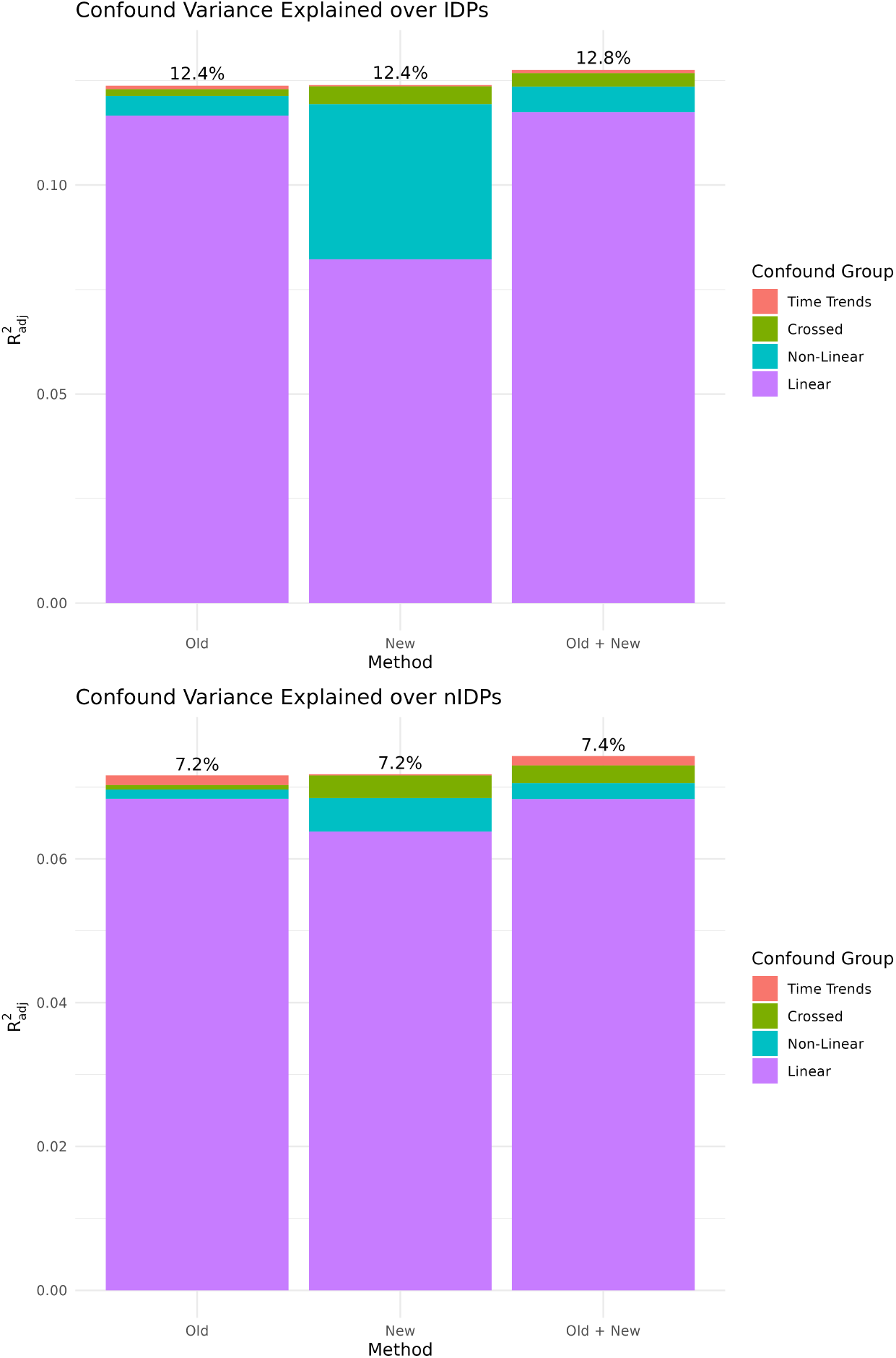
Bar plots showing the adjusted *R*^2^ for the two different confound sets and for the combined set of confounds. The variance is distributed differently between different confound groups for the two sets, but the proportion explained is very similar. Importantly, there is no notable increase in variance explained when combining the confound groups, meaning that almost all important mutual variance between confounds and IDPs/nIDPs is captured by the new confound set.

Figure 6 illustrates the proportion of variance explained in individual IDPs and nIDPs by old and new confound sets. We see very good alignment between the two confound sets across IDPs and nIDPs, with generally very small differences in the proportion of variance explained. To further verify that the two confound sets have the same effect, we also look at the correlation between IDPs/nIDPs that have been deconfounded using old and new confounds. The correlation is generally very high with the vast majority having correlations above 0.95 (98.4% for IDPs and 99.9% for nIDPs). Lastly, Figure 7 shows a Bland-Altman plot of − log_10_ p-values of univariate IDP–nIDP associations after deconfounding using new and old confounds. We see very strong agreement between p-values, especially when looking at low p-values. For 98% of IDP on nIDP associations, the two p-values are within one order of magnitude of each other. Of those p-values which are below the Bonferroni threshold controlling the Family Wise Error Rate (FWER) at 5% (*p* ≈ 10^−8^) for either old or new confounds, 54% are within one order of magnitude of each other, and 89.0% are within three orders of magnitude of each other. Two additional plots comparing the two confound sets are found in Section S1.3 of the Supplementary Material.

**Figure 6.**
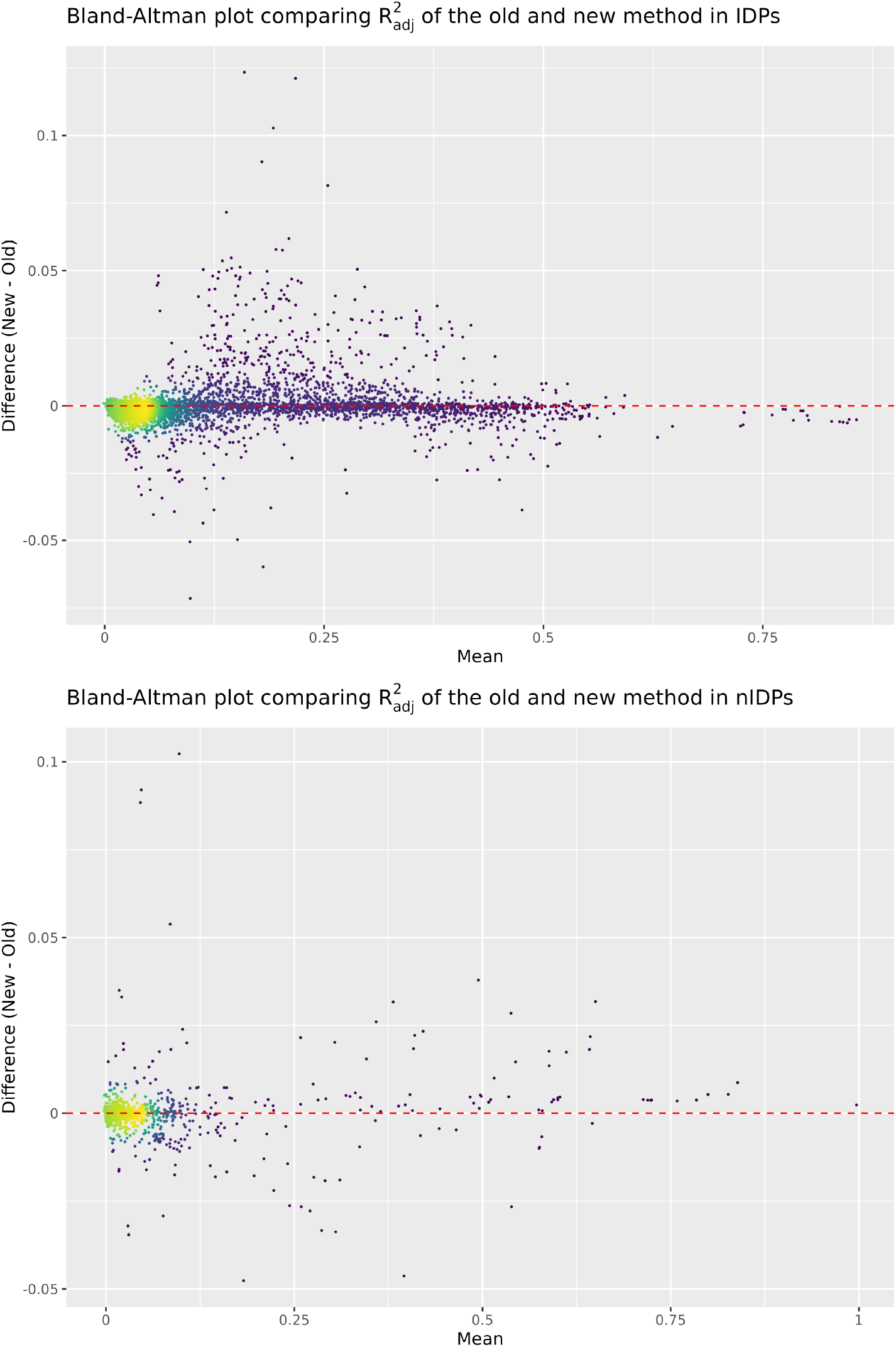
Bland-Altman plots illustrating the difference in variance explained in each IDP/nIDP by the old and new confound sets. We see that the two confound sets generally explain very similar proportions of variance in individual IDPs and nIDPs.

**Figure 7.**
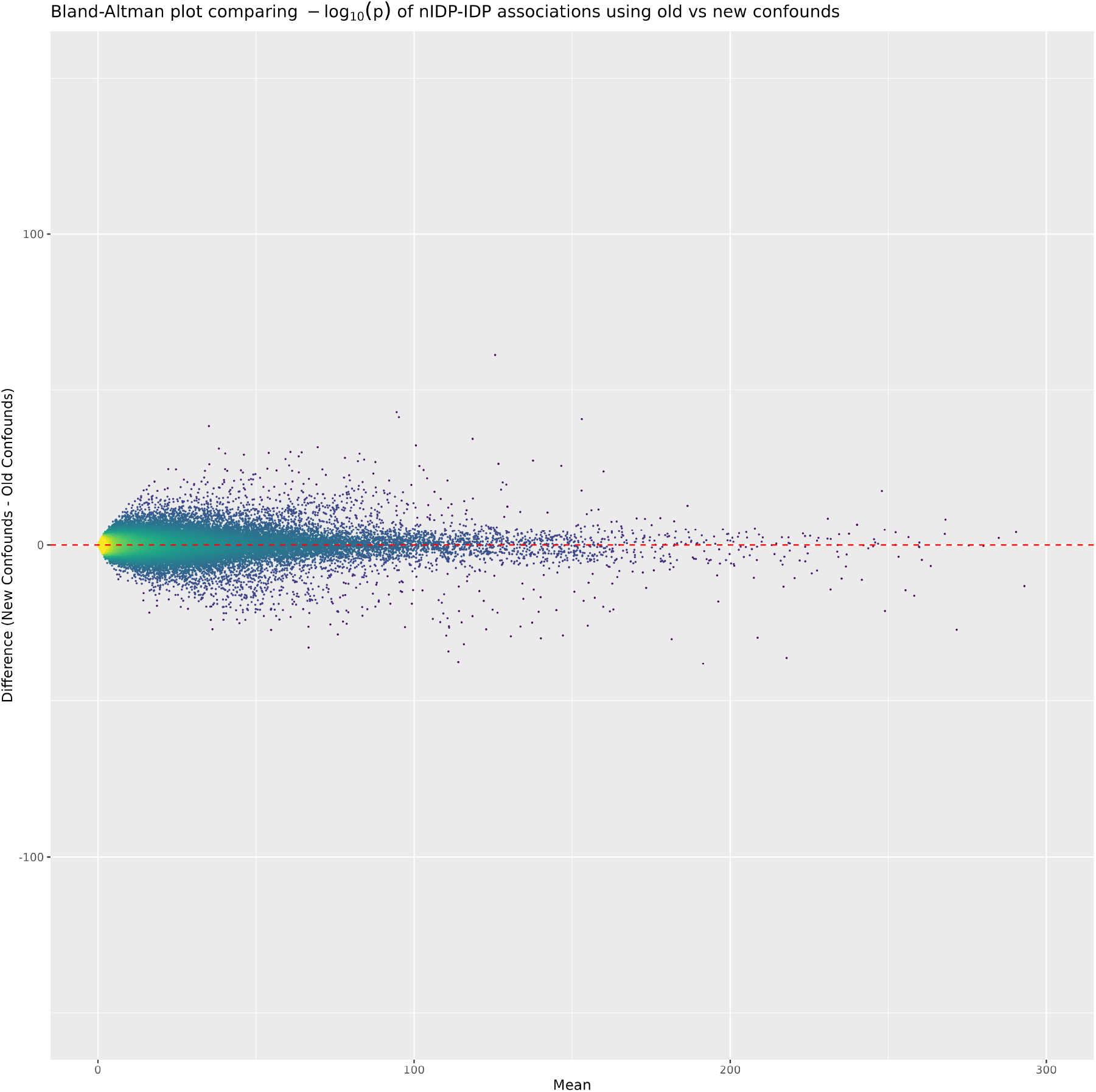
Bland-Altman plot of − log_10_(*p*) for univariate IDP on nIDP associations using old and new confounds. The plot shows very strong agreement between p-values obtained after deconfounding with either confound set.

We compared versions of the new pipeline in which single- and double-selection are used, respectively. We found very little difference between the two pipelines, with the version using double selection having a slight but noticeable advantage in explaining variance in the nIDPs. Plots comparing the two pipelines are found in the Supplementary Material, Section S1.4.

## 5 Discussion

We have proposed an improved method for confound selection on large-scale epidemiological datasets. The new confound set constructed using our pipeline consists of 312 confounds compared the previous 966 confounds, while having a very similar effect as the old set. Our proposed method is sufficiently fast to be used for forward selection (incremental forward selection of confounds, one at a time) using UK Biobank brain imaging data (tens of thousands of subjects, tens of thousands of potential confounds, and thousands of IDPs). It has the advantage of being neutral to the rate of missingness in IDPs and nIDPs, meaning that it does not systematically inflate/deflate the estimates of *R*^2^ for variables with high/low missingness. This is not the case when using uncorrected estimates of *R*^2^ on data that has been imputed, where (depending on the imputation method) higher missingness leads to inflation/deflation of the *R*^2^ estimate.

The main limitation of this work is reliance on the assumption of MCAR, which has been shown to not hold for nIDPs in general (Radosavljević et al., 2025). Future work includes developing similar methodology that only assumes Missing at Random (MAR), a weaker missingness criterion, and is therefore applicable to a larger scope of feature selection problems. Another limitation of our pipeline is that it uses classical forward selection, which has been shown to yield inflated false positive rates. There have been solutions proposed to handle this problem (Benjamini and Gavrilov, 2009, Blanchet et al., 2008, Tibshirani et al., 2016), and future work includes exploration of such options. However, in large sample settings, such as that considered by the current work, it has been shown that this problem is largely mitigated (Li et al., 2007).

The confound set produced by our pipeline is designed to be used for mass-univariate analysis of associations between IDPs and nIDPs (although the resulting confounds should be relevant for multivariate analyses too, for example, canonical correlation analysis). It therefore includes variables that explain meaningful variance in any individual IDP/nIDP. Hence, when considering a smaller problem, for example mass-univariate analysis of all IDPs on one nIDP, many of the confounds might be redundant and, consequently, inefficient power-wise. This is especially true when considering a smaller subsets of subjects. Future work therefore also includes efficient deconfounded inference when considering subsets of subjects and variables; however, as we would recommend regenerating the confound set when using a highly reduced set of subjects, that should ameliorate this concern, and the fact that the new pipeline is computationally efficient makes this easily feasible.

When regenerating confounds on a reduced subset of subjects, the choice of value for *τ*_max_ and *τ*_mean_ is an important design detail, as these thresholds prevent the inclusion of noisy confounds. We recommend the use of our thresholds for the full data set *τ*_max_ = 0.0025 and *τ*_mean_ = 0.0005 as a default starting point, but if the subset is much smaller than *n* ≈ 50000, larger thresholds might be necessary. In particular, using a smaller subset of subjects should not result in a significantly larger number of confounds, so if the pipeline returns much more than ≈ 300 confounds, it should be rerun with higher thresholds. More optimized ways of choosing *τ*_max_ and *τ*_mean_ is also an area of future inquiry.

## Data Availability

All data produced in the present work are contained in the manuscript

## Declarations

### Ethics approval and consent to participate

The UK Biobank has received ethical approval from the North West Multi-Center Research Ethics Committee (11/NW/0382).

This research project received approval from the UKB under application number 8107. This research project has adhered to the Declaration of Helsinki.

### Consent for publication

Not applicable.

### Availability of data and materials

All code can be found in the following repository: https://git.fmrib.ox.ac.uk/fsl/fastconfounds/

## Acknowledgements

The computational aspects of this research were supported by the Wellcome Trust Core Award Grant Number 203141/Z/16/Z and the NIHR Oxford BRC. The views expressed are those of the author(s) and not necessarily those of the NHS, the NIHR or the Department of Health

## 6 Supplementary Material

### S1.1 Proof of consistency of 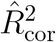

Assume the following linear model:

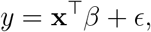

where *y* is a scalar random variable, **x** is a *d*-dimensional explanatory variable, a random vector with expectation **0** and covariance matrix Σ, *β* is a fixed *d*-dimensional vector of coefficients and *ϵ* is white noise with variance 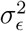. While the standard setting assumes a fixed design matrix, we consider behavior for ever-growing *n* samples of (**x**, *y*). For this linear model, the proportion of variance explained is:

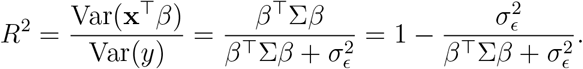

Let us assume that we have *n* = *n*_obs_ + *n*_mis_ samples of (**x**, *y*), where *n*_mis_ samples of **y** = (*y*_1_, *y*_2_, …, *y*_*n*_) have been randomly censored in an MCAR fashion at the rate of *p*_miss_. We thus construct **y**_imp_, where the missing values of **y** are imputed with 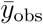, the arithmetic mean of the observed values. We now have a design matrix **X** = (**x**_1_, …, **x**_*n*_)^⊤^ and response ***y***_*imp*_ *=* (*y*_imp,1_,.., *y*_imp,*n*_)^⊤^, and we want to show that

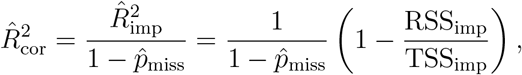

where 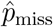 is the estimated rate of missingness, is a consistent estimator of *R*^2^.

We start by showing that

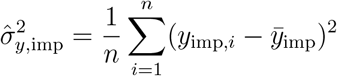

is a consistent estimator of 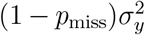. We first observe that 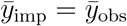. This means that 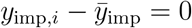 for all indices *i* where *y* is missing.

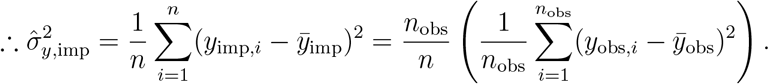

Since 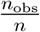 is a consistent estimator of 1 − *p*_miss_ and 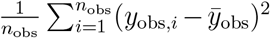 is a consistent estimator of 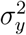 under MCAR, it follows from the continuous mapping theorem that 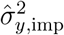 is a consistent estimator of 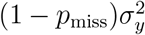.

We continue by proving that

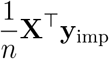

is a consistent estimator of (1 − *p*_miss_)Cov(**x**, *y*) = (1 − *p*_miss_)Σ*β*. We know that

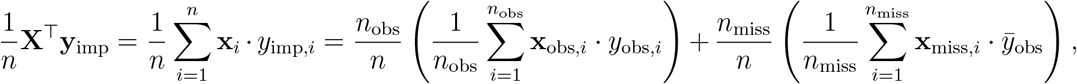

and since 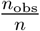 is a consistent estimator of 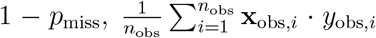 is a consistent estimator of Cov(**x**, *y*) and 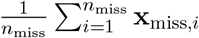 is a consistent estimator of 𝔼 (**x**) = **0**, it follows from the continuous mapping theorem that 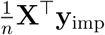 is a consistent estimator of (1 − *p*_miss_)Cov(**x**, *y*) = (1 − *p*_miss_)Σ*β*.

Now we want to prove that

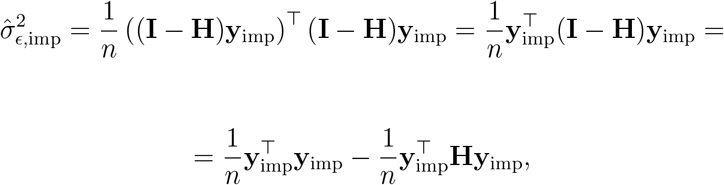

where **H** = **X**(**X**^⊤^**X**)^−1^**X**^⊤^ is a consistent estimator of 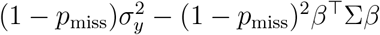.

We know that

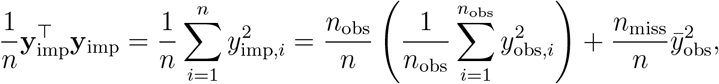

and since 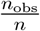 is a consistent estimator of 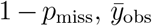 is a consistant estimator of 𝔼 (*y*) = 0 and 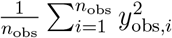 is a consistent estimator of 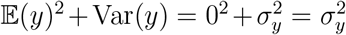, it follows from the continuous mapping theorem that 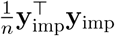 is a consistent estimator of 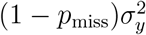.

We continue by observing that

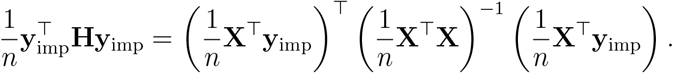

Here we use our previous result that 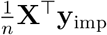 is a consistent estimator of (1 − *p*_miss_)Σ*β*, as well as the fact that 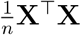 is a consistent estimator of Σ, and get from the continuous mapping theorem that 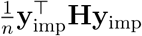 is a consistent estimator of

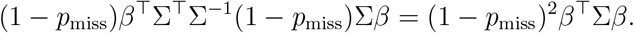

From this, it finally follows that 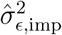 is a consistent estimator of

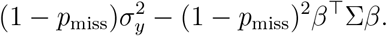

We now have all results necessary to prove the consistency of 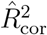. We note that

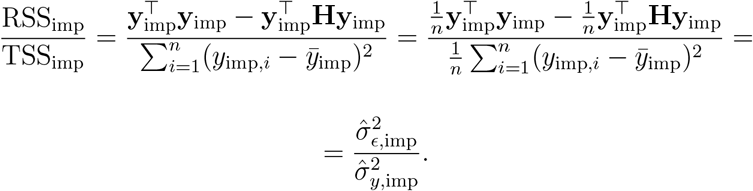

It thus follows that

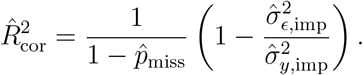

Using our results and a combination of the continuous mapping theorem and Slutsky’s theorem, we finally get that 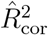 is a consistent estimator of

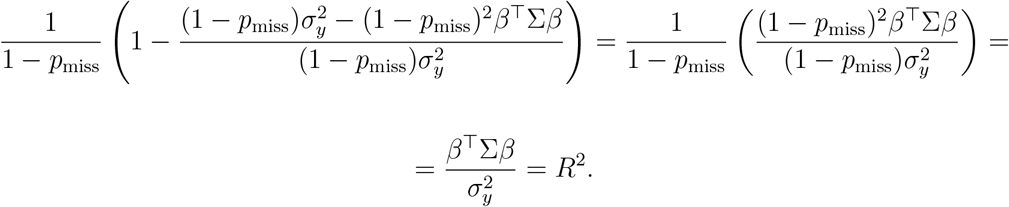

### S1.2 Algorithm

The proposed forward selection algorithm has two pre-determined thresholds *τ*_mean_ and *τ*_max_ which determine when no remaining potential confounds are important enough to be included in the final confound set. The algorithm is described in detail below using pseudocode. The functions `P_MISS`, `PINV`and `MEAN_IMPUTER` perform the calculation of column-wise missingness rates, the Moore-Penrose pseudo-inverse and mean imputation respectively. All other function names are assumed to be self-explanatory. The operator * is used for matrix multiplication, all other operations are assumed to be applied element-wise. All data is standardised before running the algorithm.

We can see from the algorithm that the most computationally costly operations is initially calculating the pseudo-inverse of **C**_fix_, and also performing the matrix multiplication 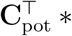 IDPs in each iteration. If *d*_fix_ is the number of fixed confounds, *d*_pot_ the number of potential confounds, *d*_IDPs_ the number of IDPs, *n* the number of subjects, and *s*_pot_ *<< d*_pot_ the number of relevant potential IDPs, then the time complexity of our algorithm is:

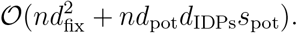

It should also be noted that all computations are vectorised as we only use matrix multiplication and element-wise matrix/vector manipulation in the algorithm.

#### Algorithm 1

Selecting Potential Confounds

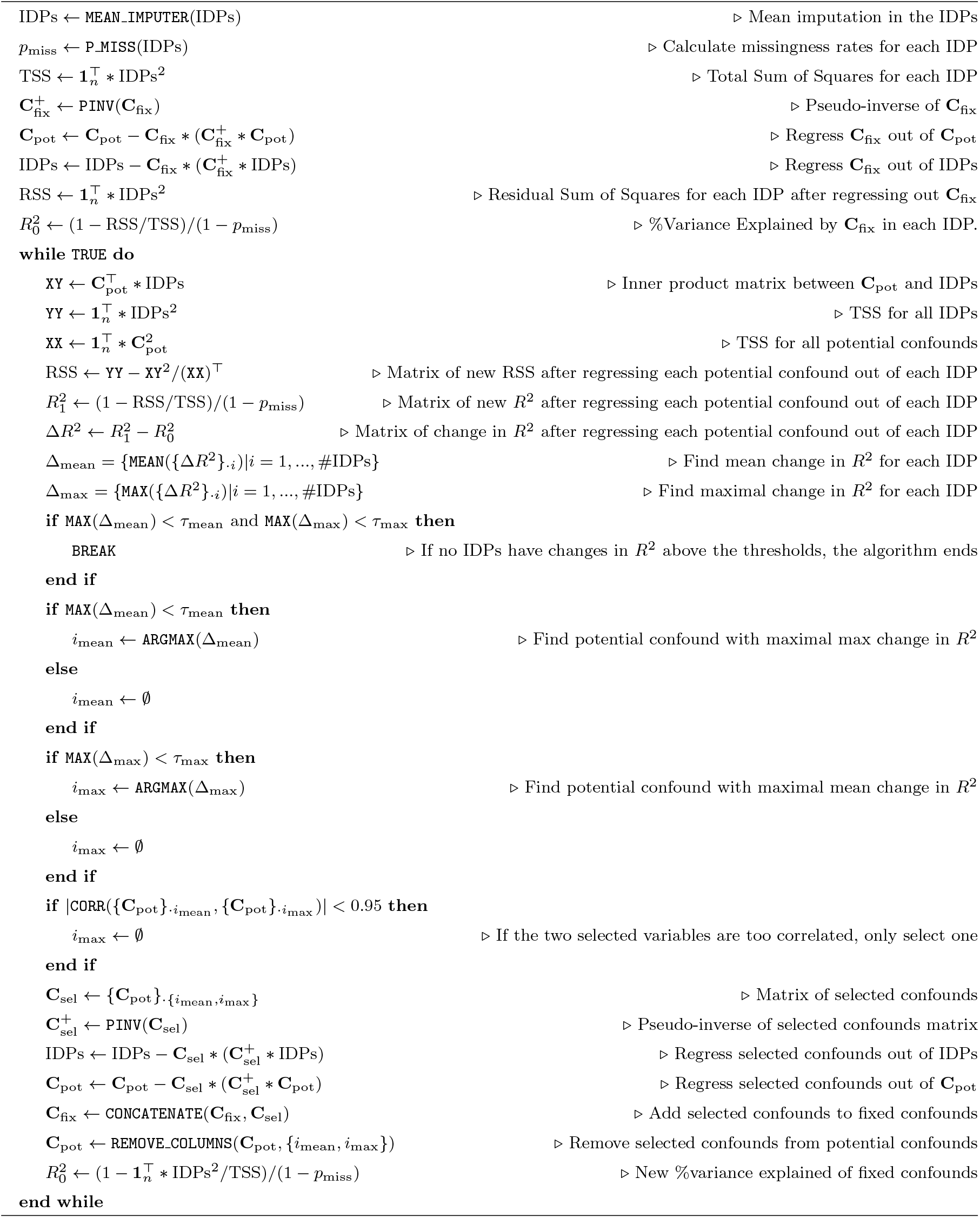

### S1.3 Further Comparisons of Old and New Confounds

**Figure S1.**
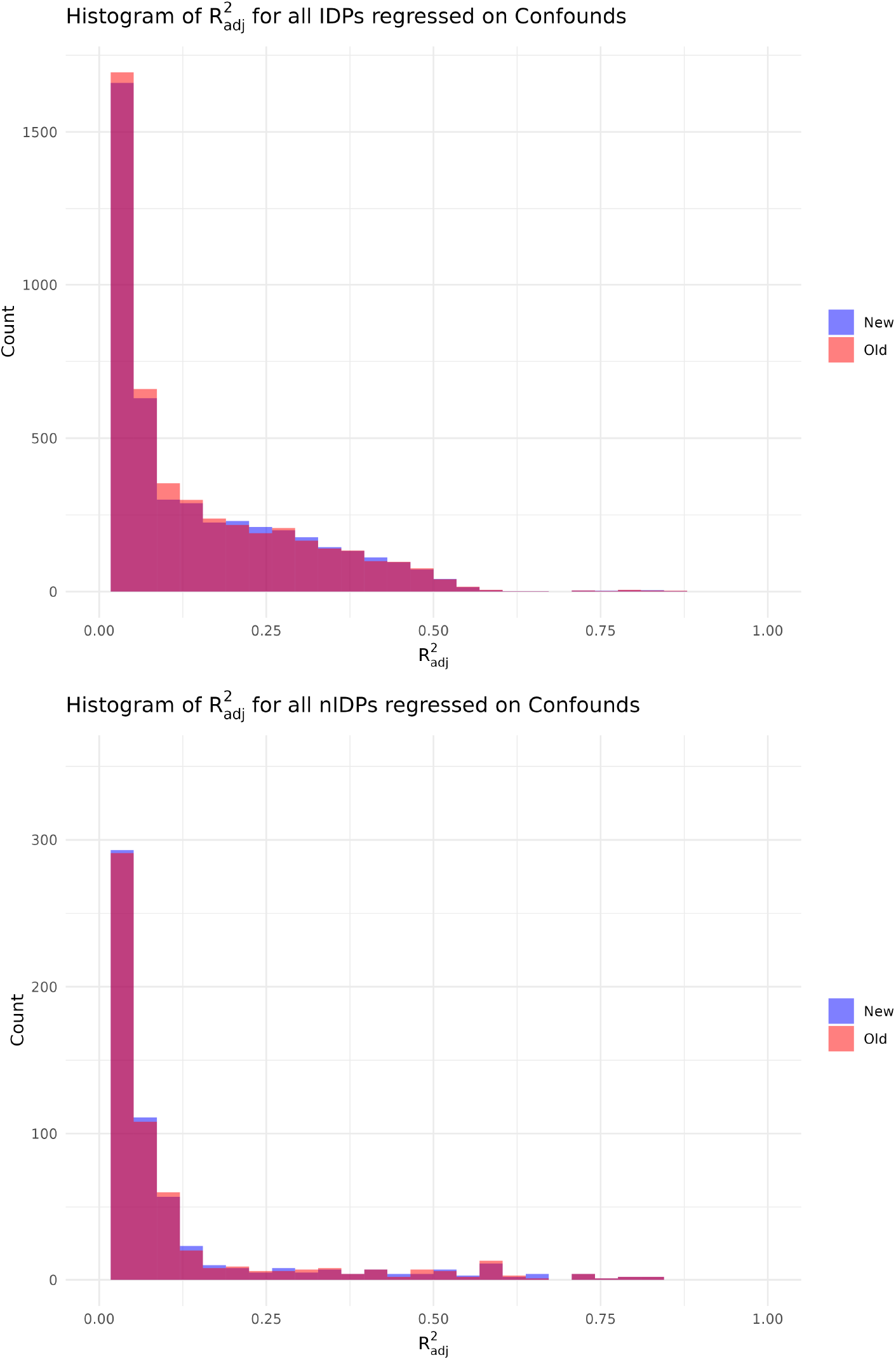
Overlapping histograms of the proportion of variance explained in the IDPs and nIDPs by old and new confounds. We see that there is great overlap in distribution of variance explained for the old and new confounds.

**Figure S2.**
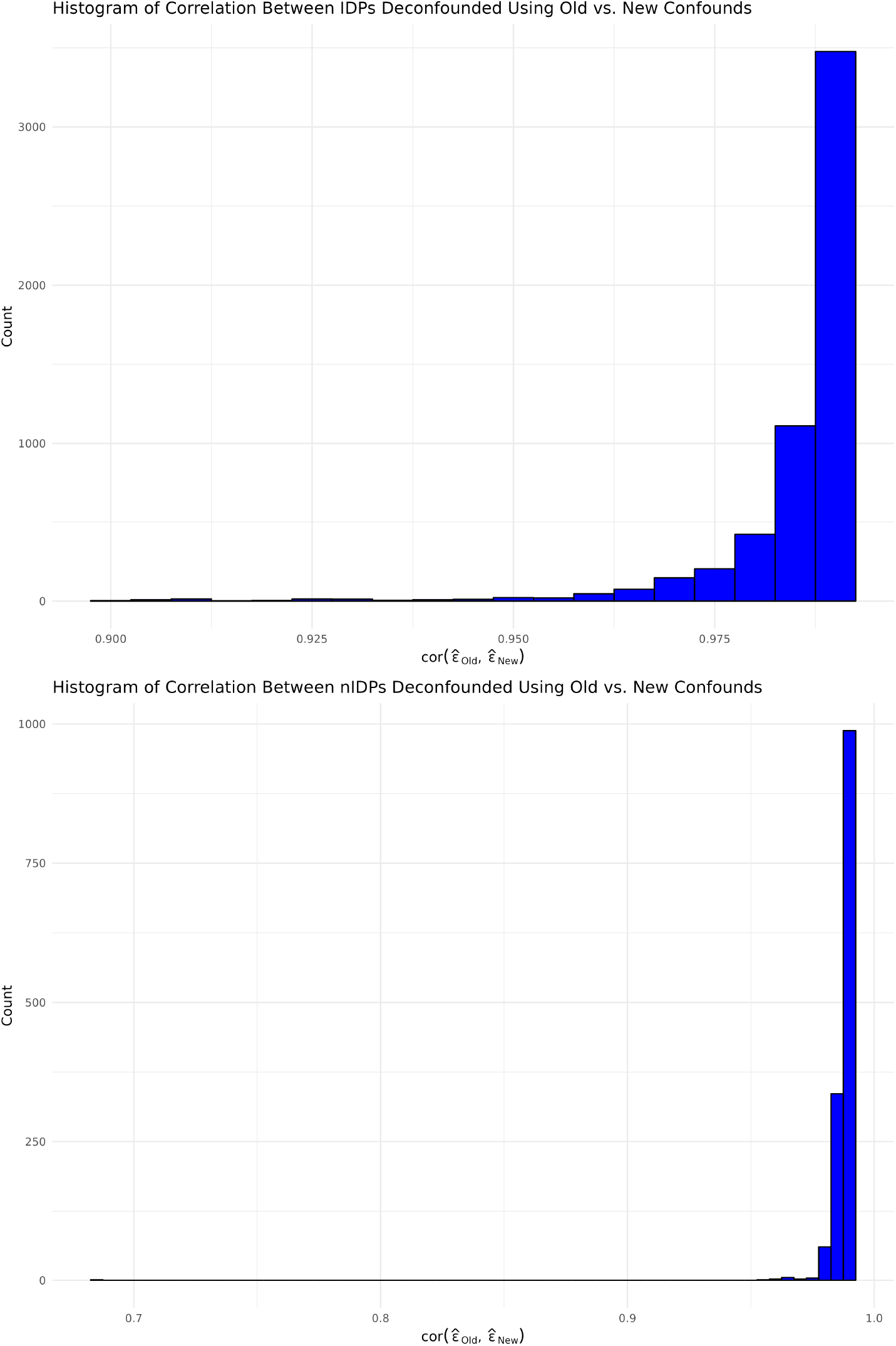
Histograms of the correlation between IDPs and nIDPs deconfounded using old and new confounds. We see a very high correlation between IDPs and nIDPs deconfounded using different confound sets, meaning that deconfounding has a very similar effect on IDPs/nIDPs when using either confound set.

### S1.4 Comparing Double Selection and Single Selection

**Figure S3.**
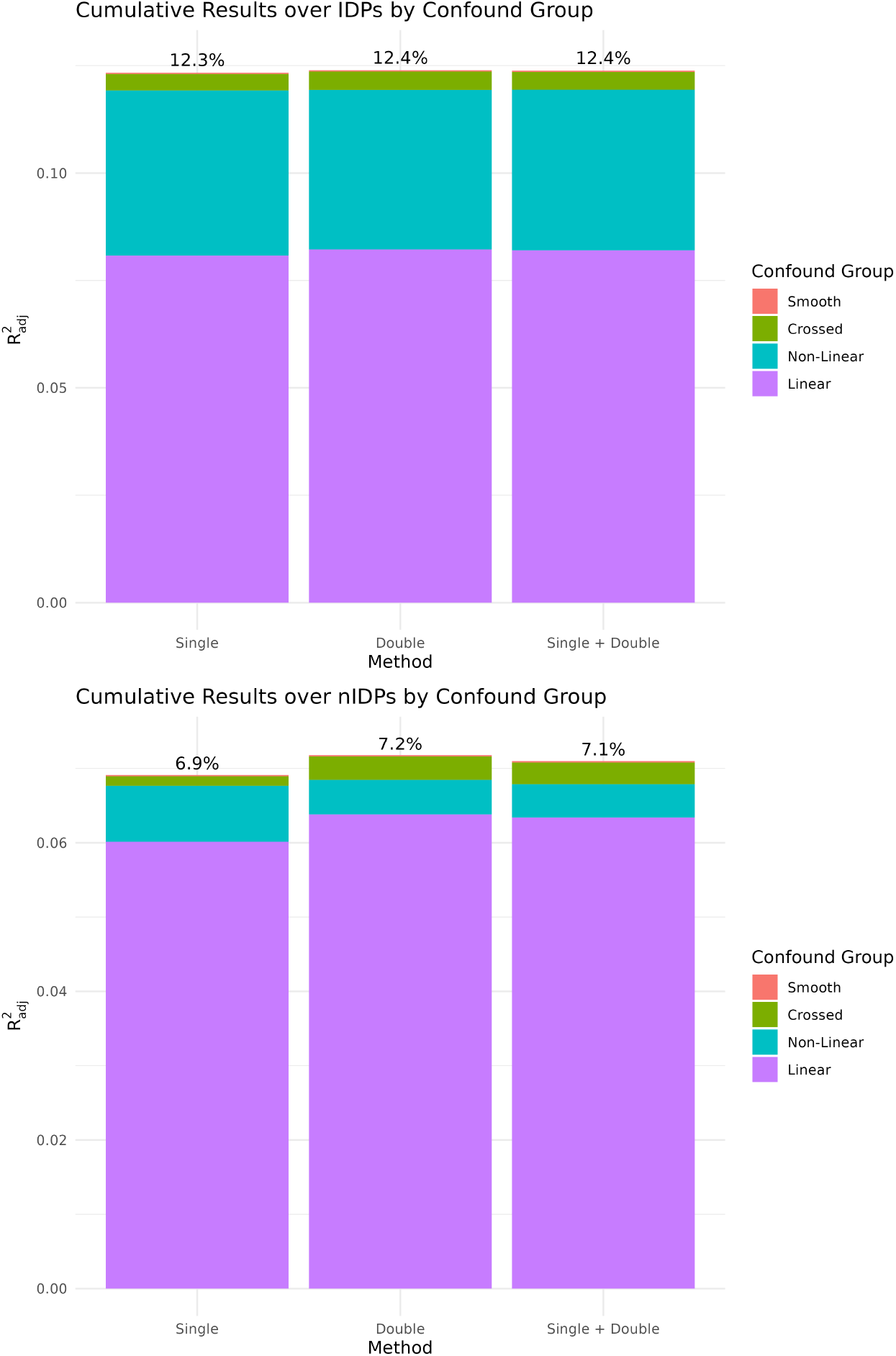
Bar plots comparing the average variance explained using double-vs. using single-selection. For IDPs, the proportions are almost identical. For nIDPs, double selection leads to a small but noticeable rise in variance explained.

**Figure S4.**
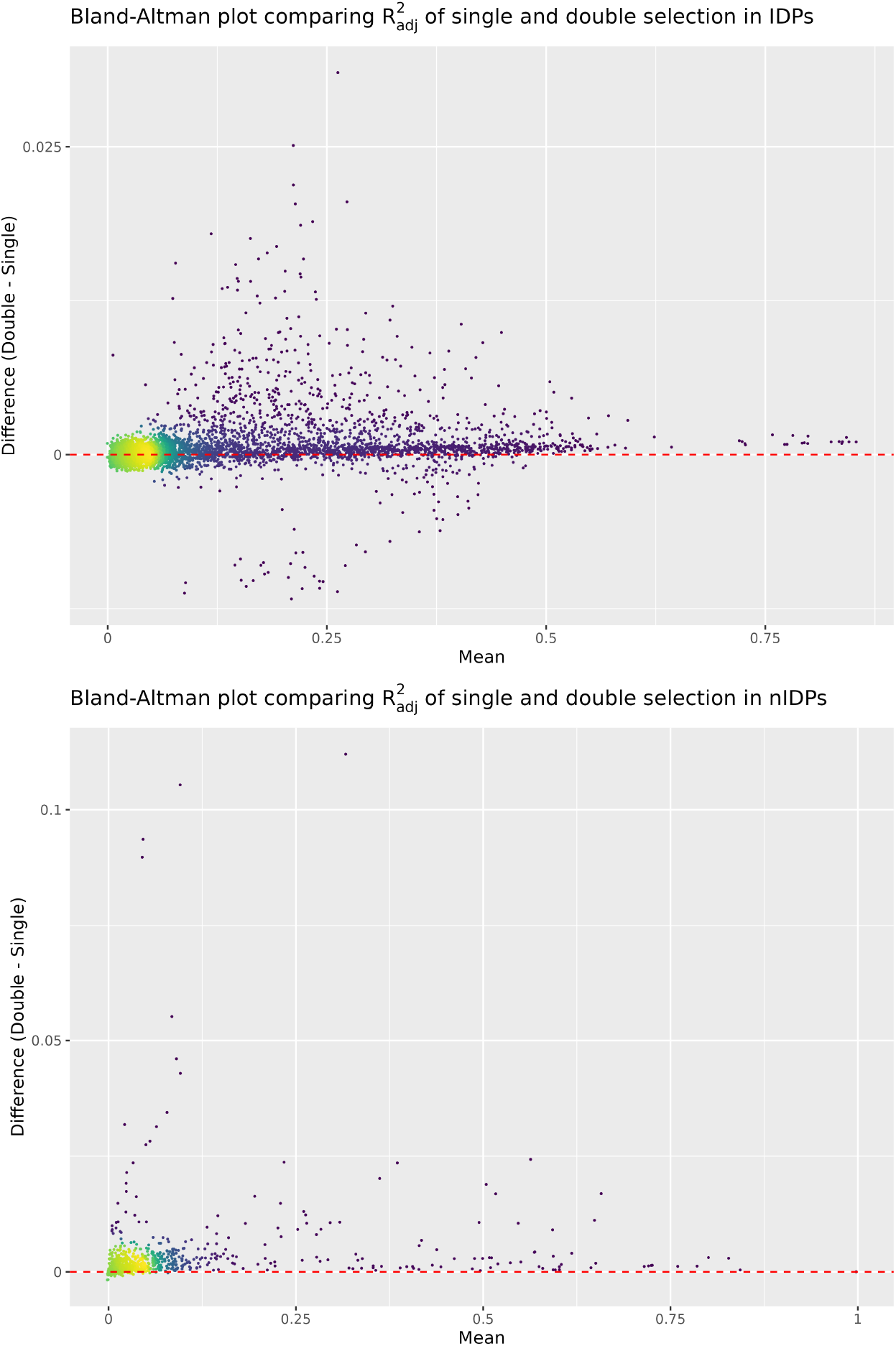
Bland-Altman plots illustrating the difference in variance explained in each IDP/nIDP by confound sets constructed using double-vs. single selection. For IDPs, we see very strong agreement between the two different methods. For nIDPs, we see that double-selection leads to consistently higher variance explained but generally not drastically higher.

**Figure S5.**
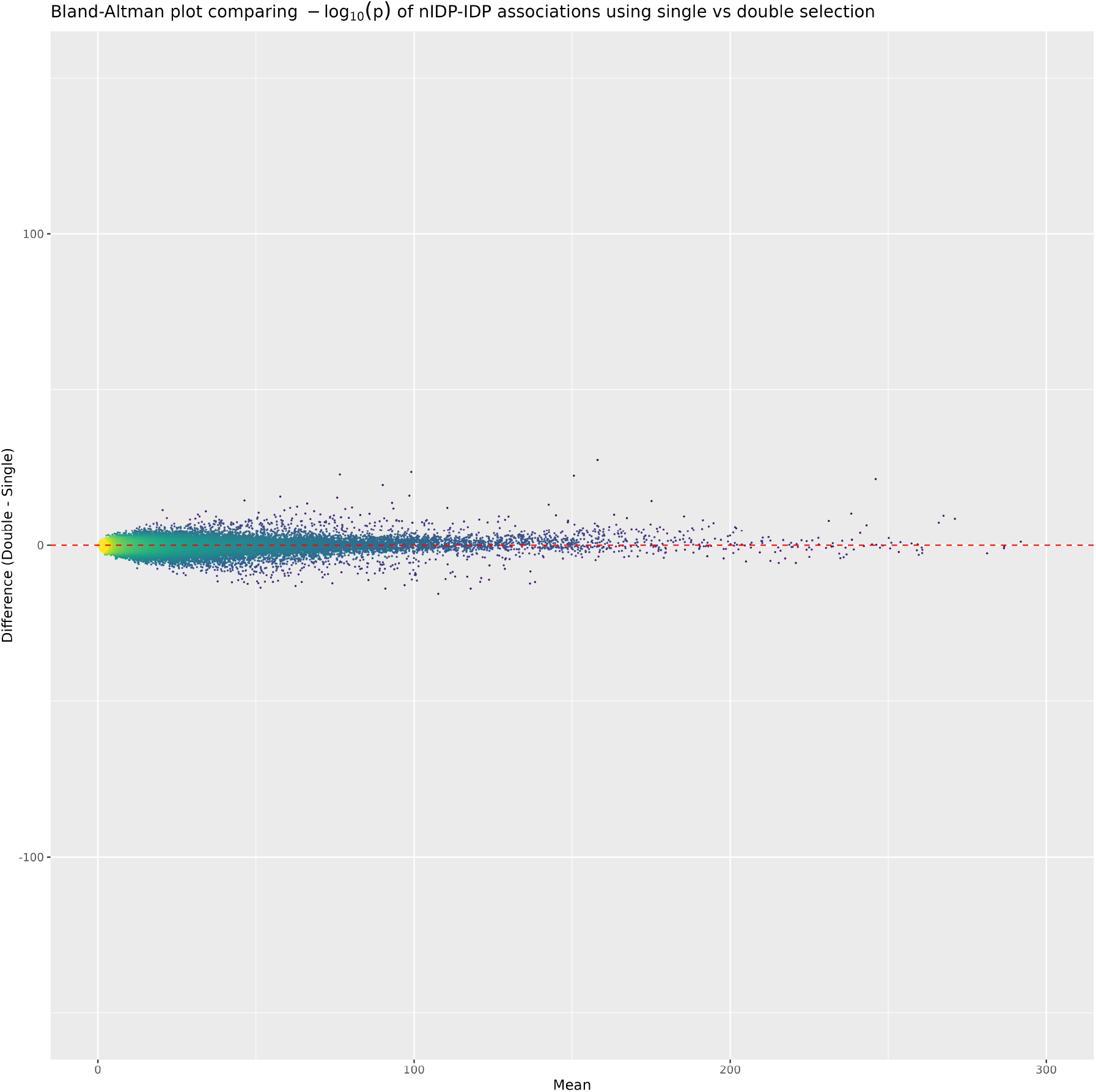
Bland-Altman plot of − log_10_(*p*) for univariate IDP on nIDP associations using confounds constructed with single- or double-selection. The plot shows that p-values obtained after deconfounding with either confound set are extremely close.

